# Time is of the essence: containment of the SARS-CoV-2 epidemic in Switzerland from February to May 2020

**DOI:** 10.1101/2020.07.21.20158014

**Authors:** Christian L. Althaus, Daniel Probst, Anthony Hauser, Julien Riou

**Affiliations:** Institute of Social and Preventive Medicine, University of Bern, Bern, Switzerland; Department of Chemistry and Biochemistry, National Center for Competence in Research NCCR TransCure, University of Bern, Bern, Switzerland

## Abstract

**AIM:** In late February and early March 2020, Switzerland experienced rapid growth of severe acute respiratory syndrome coronavirus 2 (SARS-CoV-2) infections with 30,243 confirmed cases and 1,860 deaths as of 10 May 2020. The sequential introduction of non-pharmaceutical interventions (NPIs) resulted in successful containment of the epidemic. A better understanding of how the timing of implementing NPIs influences the dynamics and outcome of SARS-CoV-2 epidemics will be crucial for the management of a potential resurgence in Switzerland.

**METHODS:** We developed a dynamic transmission model that describes infection, hospitalization, recovery and death due to SARS-CoV-2 in Switzerland. Using a maximum likelihood framework, we fitted the model to aggregated daily numbers of hospitalized patients, ICU occupancy and death from 25 February to 10 May 2020. We estimated critical parameters of SARS-CoV-2 transmission in Switzerland and explored counterfactual scenarios of an earlier and later implementation of NPIs.

**RESULTS:** We estimated the basic reproduction number *R*_0_ = 2.61 (95% compatibility interval, CI: 2.51–2.71) during the early exponential phase of the SARS-CoV-2 epidemic in Switzerland. After the implementation of NPIs, the effective reproduction number approached *R*_*e*_ = 0.64 (95% CI: 0.61–0.66). Based on the observed doubling times of the epidemic before and after the implementation of NPIs, we estimated that one week of early exponential spread required 3.1 weeks (95% CI: 2.8–3.3 weeks) of ‘lockdown’ to reduce the number of infections to the same level. Introducing the same sequence of NPIs one week earlier or later would have resulted in substantially lower (399, 95% prediction interval, PI: 347–458) and higher (8,683, 95% PI: 8,038–9,453) numbers of deaths, respectively.

**CONCLUSIONS:** The introduction of NPIs in March 2020 prevented thousands of SARS-CoV-2-related deaths in Switzerland. Early implementation of NPIs during SARS-CoV-2 outbreaks can reduce the number of deaths and the necessary duration of strict control measures considerably.

---

> *“That’s a window of opportunity so I’m reminding; there is time, the time is ticking and time is of the essence in this outbreak*.*”*
>
> – Director-General of the World Health Organization, 11 February 2020

## INTRODUCTION

Switzerland reported its first confirmed case of severe acute respiratory syndrome coronavirus 2 (SARS-CoV-2) on 25 February 2020 [1]. The first case was diagnosed in the canton of Ticino, which borders the Italian region of Lombardy that already experienced an ongoing SARS-CoV-2 outbreak at the time. In the subsequent days, additional cases were reported in several cantons leading to a rapid spread of SARS-CoV-2 all over Switzerland. Starting on 28 February 2020, the federal government sequentially issued a number of non-pharmaceutical interventions (NPIs) that included closing of primary schools, non-essential shops, and restaurants, culminating in the prohibition of gatherings of more than five people on 20 March 2020 (‘lockdown’). The SARS-CoV-2 epidemic subsequently decreased with 30,243 confirmed cases and 1,860 deaths until 10 May 2020 [2] when major restrictions with regards to schools, non-essential shops, and restaurants were lifted again. In order to better prepare for a resurgence of cases in Switzerland, it is crucial to understand how the timing of implementing NPIs influences the dynamics and outcome of SARS-CoV-2 epidemics.

Mathematical and statistical modeling has been influential in the analysis of the SARS-CoV-2 pandemic. A number of key studies provided early estimates of the basic reproduction number *R*_0_ [3, 4], analyzed the dynamics and control of the outbreak in Wuhan, China [5], investigated the feasibility of outbreak control by case isolation [6], and estimated the infection fatality ratio (IFR) [7–10]. For Switzerland, Lemaitre et al. [11] assessed the impact of non-pharmaceutical interventions on SARS-CoV-2 transmission for various cantons (states). Another study estimated that 54,000 (95% credible interval: 36,000–73,000) deaths could have occurred in Switzerland assuming no interventions had taken place at all [12]. To date, there are no studies that investigate the effects of counterfactual scenarios for Switzerland, with interventions being implemented at different time points, on the expected number of infections, hospitalized cases and deaths.

We developed a SARS-CoV-2 transmission model for Switzerland that describes the progression of patients through hospitals and intensive care units (ICUs) to recovery or death. We fitted the model to aggregated data on the number of hospitalized cases, ICU occupancy and deaths using a maximum likelihood framework. We estimated *R*_0_ during the early epidemic spread and the effective reproduction number *R*_*e*_ after the implementation of NPIs. Furthermore, we explored the impact of an earlier and a later introduction of control measures on SARS-CoV-2-related morbidity and mortality in Switzerland.

## METHODS

### Transmission model

We developed a deterministic, population-based transmission model for SARS-CoV-2 in Switzerland. The model includes within-hospital dynamics and is described by the following set of ordinary differential equations (ODEs) (fig. 1):

**Figure 1.**
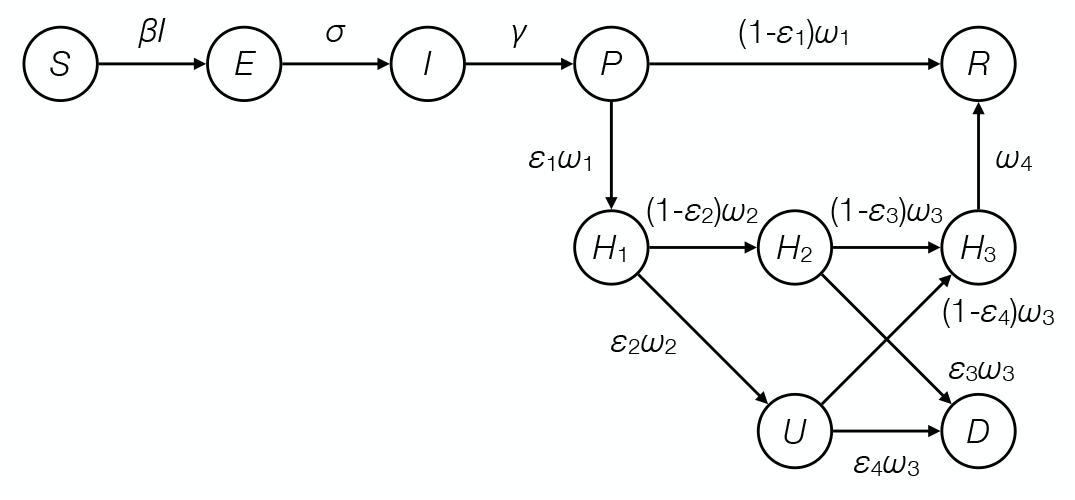
Schematic illustration of the SARS-CoV-2 transmission model. After infection, susceptible individuals (*S*) get exposed (*E*) before they become infectious (*I*). The infection then progresses (*P*) and individuals are either admitted to hospital (*H*_1_) or recover (*R*). At the hospital, some patients enter the intensive care unit (ICU, *U*) where they either die (*D*) or go on to recover in the hospital (*H*_3_). Hospitalized patients can also die outside of ICU (*H*_2_).

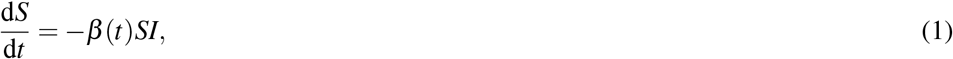

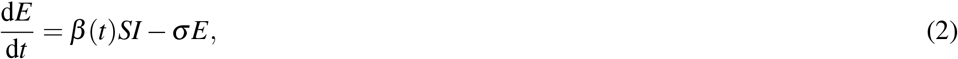

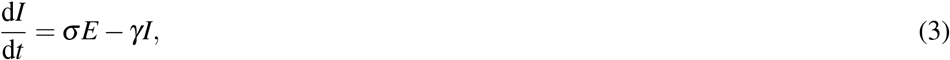

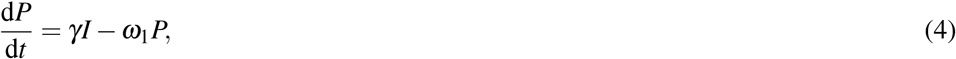

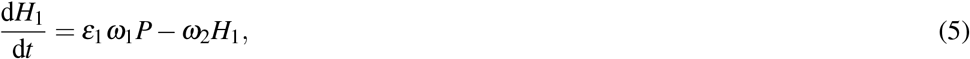

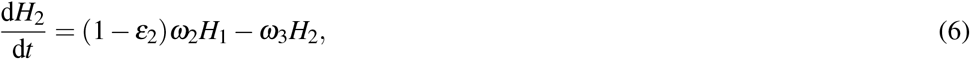

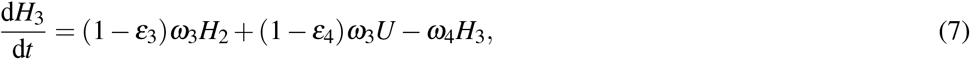

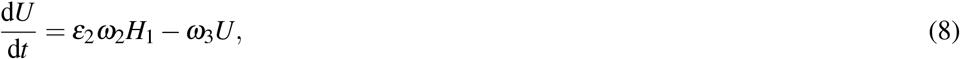

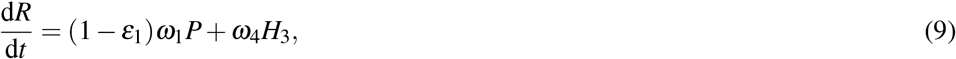

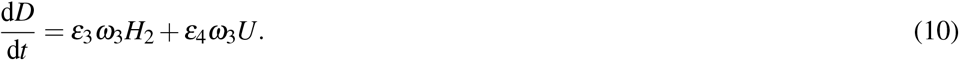

Susceptible individuals *S* get infected by infectious individuals *I* at rate *β*. Exposed individuals *E* move through an incubation period at rate *σ* before they become infectious for 1/*γ* days. The infection then progresses (*P*) and a fraction *ε*_1_ of individuals are admitted to hospital (*H*_1_) while the others recover (*R*) at rate *ω*_1_. Within the hospital, a fraction *ε*_2_ of patients enter ICU (*U*) after 1/*ω*_2_ days while the others remain in hospital (*H*_2_). Patients either die in ICU (*U*) or hospital beds (*H*_2_) with probabilities *ε*_3_ and *ε*_4_, or remain in hospital (*H*_3_) for another 1/*ω*_4_ days and recover (*R*). For simplicity, we do not explicitly consider deaths outside hospitals. Hence, the probabilities of death in hospital bed (*ε*_3_) and ICU (*ε*_4_) also account for deaths outside hospitals (table 1).

**Table 1.**
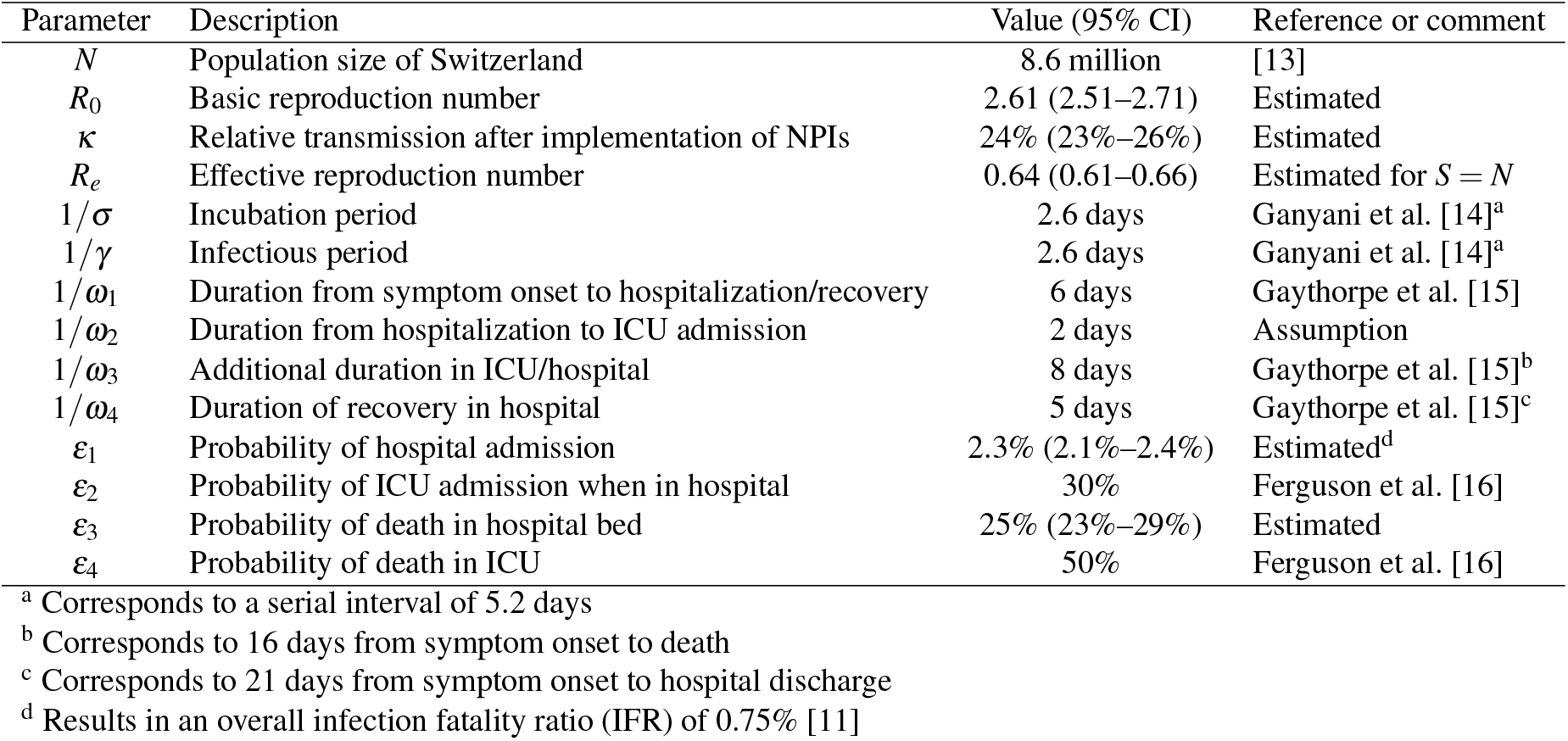
Parameters of the SARS-CoV-2 transmission model.

### Non-pharmaceutical interventions (NPIs)

The Swiss government issued a number of NPIs in response to the SARS-CoV-2 epidemic. On 28 February 2020, the federal government declared a ‘special situation’ according to the Epidemics Act and banned events of more than 1,000 people. Later, the government issued an information campaign starting on 1 March 2020. On 13 March 2020, the ban on events was strengthened with a new limit of 100 people. Closure of primary schools was ordered on 14 March 2020. The federal government moved to an ‘extraordinary situation’ according to the Epidemics Act on 16 March 2020 and closed non-essential shops and restaurants by 17 March 2020. Finally, gatherings of more than five people were prohibited on 20 March 2020 (‘lockdown’).

The precise effects of the different interventions on reducing the transmission rate and *R*_*e*_ are challenging to quantify [12], but Lemaitre et al. [11] showed that NPIs in Switzerland led to a strong reduction of *R*_*e*_ starting around 7 March 2020 for a period of about two weeks. In order to capture this reduction in *R*_*e*_, we assumed that the reduction in the transmission rate *β* (*t*) follows a logistic function:

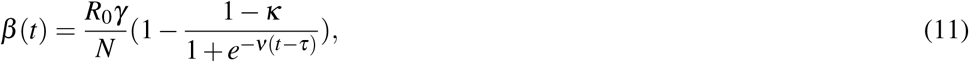

where *κ* is the relative transmission that is being approached after implementing all NPIs, *τ* is the midpoint of the reduction in transmission, and *ν* is the slope of the sigmoid function. We chose *ν* = 0.5 which roughly corresponds to a reduction in transmission over a period of two weeks. We set *τ* to 17 March 2020 which provided the best fit of the model to the data. We assumed that the transmission rate did not increase until 10 May 2020. We also calculated 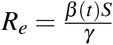

### Data

We used publicly available data on daily numbers of confirmed SARS-CoV-2 cases, hospitalized patients, ICU occupancy, and deaths in Switzerland [2]. The data set consists of aggregated cantonal numbers that are curated and provided by OpenZH, the specialist unit for open government data from the canton of Zurich [17]. We considered the time period from the report of the first confirmed case (25 February 2020) until the last day before major NPIs were lifted (10 May 2020).

### Maximum likelihood estimation

The transmission model was initialized with *I*(0) = 1 at *ι* days before the report of the first case. We then numerically integrated the ODEs using the package deSolve [18] for the R software environment for statistical computing [19]. We adapted a previously used maximum likelihood framework [20, 21] to fit the model to daily numbers of hospitalized patients, ICU occupancy and death. The likelihoods to observe the daily change in hospitalizations, Δℋ (*t*), and the daily change in ICU occupancy, Δ𝒰 (*t*), on day *t* can be described by two Skellam distributions:

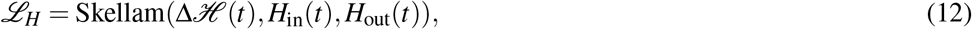

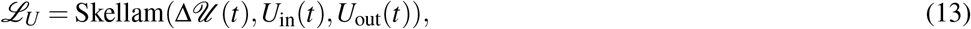

Where

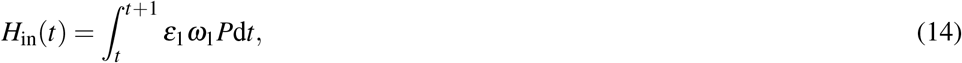

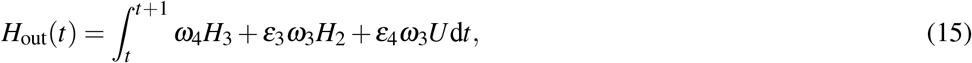

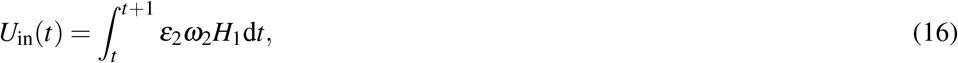

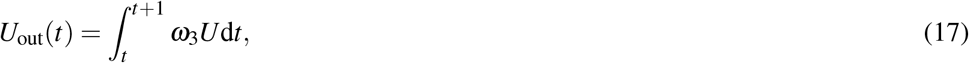

are the modeled numbers of patients that enter and leave the hospital and ICU on day *t*, respectively. The observed daily numbers of deaths Δ𝒟 (*t*) were assumed to be Poisson distributed:

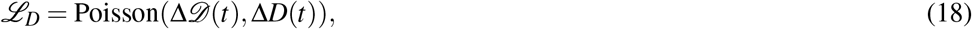

Where

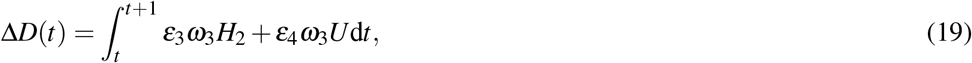

is the modeled incidence of death on day *t*. We then minimized the sum of the negative log-likelihoods to derive maximum likelihood estimates of the following model parameters: the basic reproduction number *R*_0_, the relative transmission after implementation of NPIs *κ*, the probability of hospital admission *ε*_1_, the probability of death in hospital bed *ε*_3_, and the time *ι* at which the model was initialized.

95% compatibility intervals (CIs) [22] of parameter estimates were obtained via the likelihood profile (*R*_0_ *κ, ε*_1_, *ε*_3_) or from 10,000 bootstrap samples making use of the asymptotic normality of the maximum likelihood estimates (*R*_*e*_ and doubling times). Using these bootstrapped parameter sets, we also derived prediction intervals (PIs) for the model curves by simulating Poisson-distributed daily incidences of infections and deaths and Skellam-distributed changes in the numbers of hospitalized patients and ICU occupancy for each epidemic trajectory. We then used the 2.5% and 97.5% quantiles from the simulated trajectories at each time point *t* to construct point-wise intervals.

Epidemic doubling times were calculated from *R*_0_ and *R*_*e*_ using the generation time distribution from the transmission model as described in Wallinga et al. [23]. All data and R code files are available on GitHub: https://github.com/calthaus/swiss-covid-epidemic.

### Counterfactual scenarios

We simulated counterfactual scenarios to explore the effects of an earlier and later introduction of NPIs. To this end, we shifted the sigmoid reduction in the transmission rate *β* by *n* days. We then ran the model with the maximum likelihood estimates and 1,000 bootstrapped parameter combinations. We calculated the expected number of SARS-CoV-2 infections, hospitalized patients, ICU admissions, and deaths assuming that NPIs continued to reduce the number of new infections after 10 May 2020.

## RESULTS

The transmission model fitted the daily data on hospitalized patients, ICU occupancy, and deaths during the SARS-CoV-2 epidemic in Switzerland well (fig. 2). In February and early March 2020, the daily number of new infections increased exponentially with a basic reproduction number *R*_0_ = 2.61 (95% CI: 2.51–2.71). The sequential introduction of NPIs reduced the transmission rate to *κ* = 24% (95% CI: 23%–26%), resulting in a effective reproduction number *R*_*e*_ = 0.64 (95% CI: 0.61–0.66) (fig. 3). The values of *R*_0_ and *R*_*e*_ correspond to an epidemic doubling time of 2.9 days (95% CI: 2.8–3.1 days) during the early phase of the epidemic and a half-life of 8.9 days (95% CI: 8.3–9.7 days) after implementation of all NPIs. Hence, one week of exponential increase in new infections during the early epidemic spread required 3.1 weeks (95% CI: 2.8–3.3 weeks) of ‘lockdown’ to reduce the number of infections to the same level.

**Figure 2.**
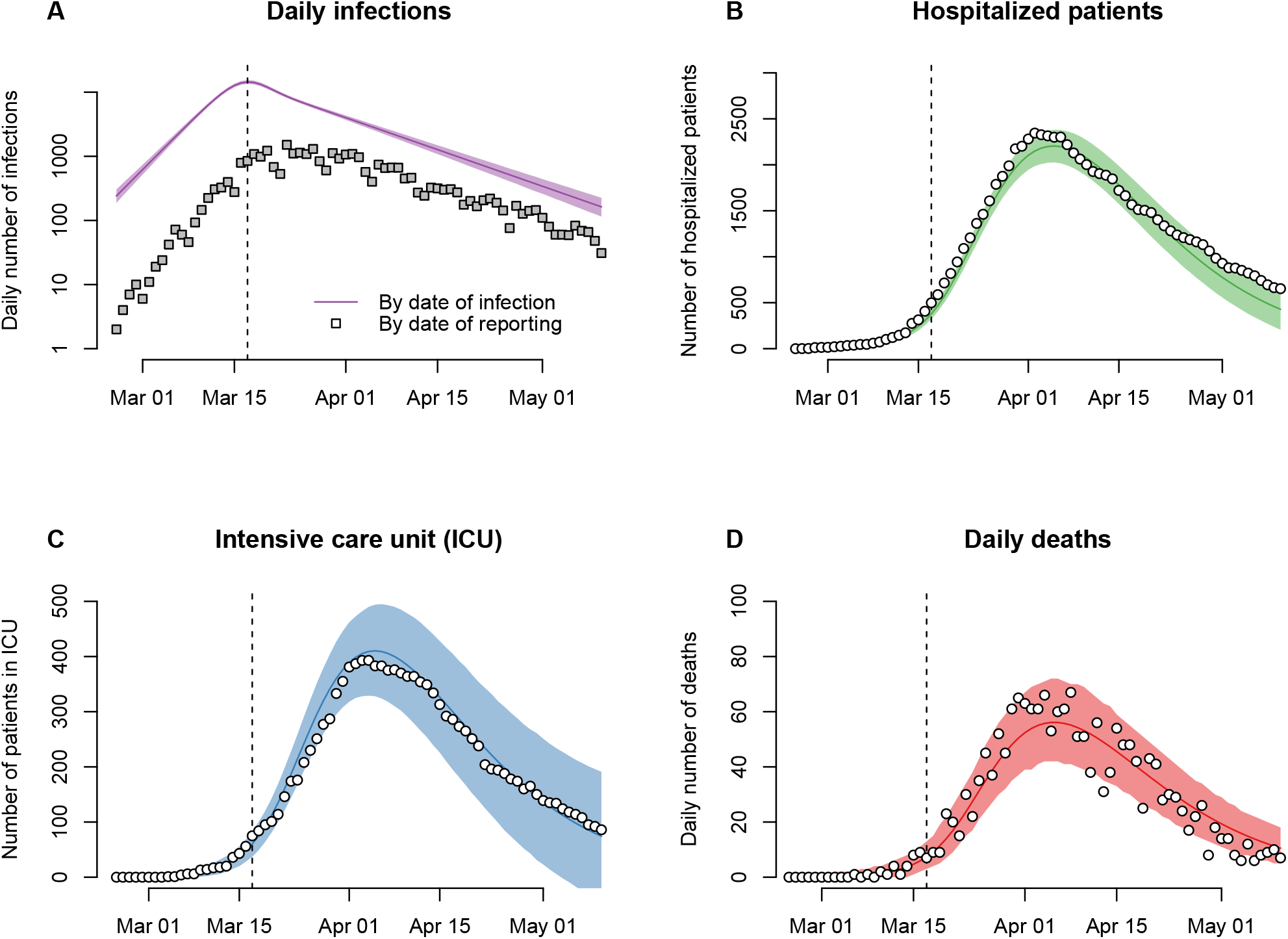
Modeled number of new infections, hospitalized patients, intensive care unit (ICU) occupancy, and deaths during the SARS-CoV-2 epidemic in Switzerland. The solid lines show the maximum likelihood estimate of the model and the shaded areas correspond to the 95% prediction intervals. The model was fitted to the data shown as white circles. Reported number of infections (gray squares) are shown for comparison. The vertical dashed line indicates the strengthening of social distancing on 17 March 2020.

**Figure 3.**
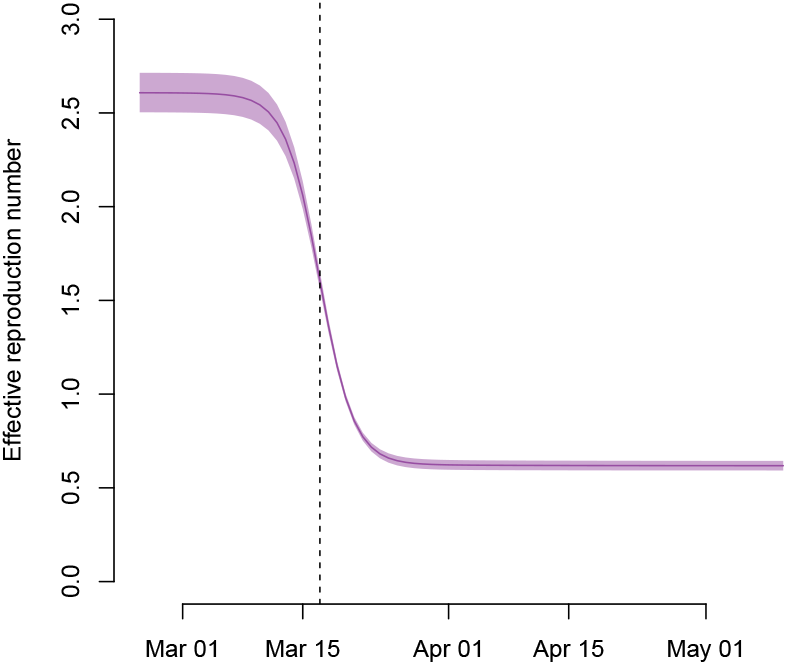
Reduction of the effective reproduction number *R*_*e*_ during the SARS-CoV-2 epidemic in Switzerland. The solid line shows the maximum likelihood estimate of the model and the shaded area corresponds to the 95% compatibility interval. We assumed that the sequential introduction of NPIs resulted in a sigmoid reduction of the transmission rate over a period of around 2 weeks. The vertical dashed line indicates the strengthening of social distancing on 17 March 2020.

The cumulative number of SARS-CoV-2 infections in Switzerland reached 264,287 (95% PI: 251,776–276,430) by 10 May 2020, corresponding to an infection attack rate of 3.1% (95% PI: 2.9%–3.2%) for Switzerland overall. The diagnosed and reported infections represent only a fraction of the overall epidemic in Switzerland. Based on the assumed IFR of 0.75%, we estimated that the proportion of new infections that were diagnosed and reported at the beginning of the epidemic was around 10% and increased to around 20% before the strengthening of social distancing measures on 17 March 2020 (fig. 4). During the ‘lockdown’, the proportion of diagnosed and reported infections decreased and stabilized at around 10% again.

**Figure 4.**
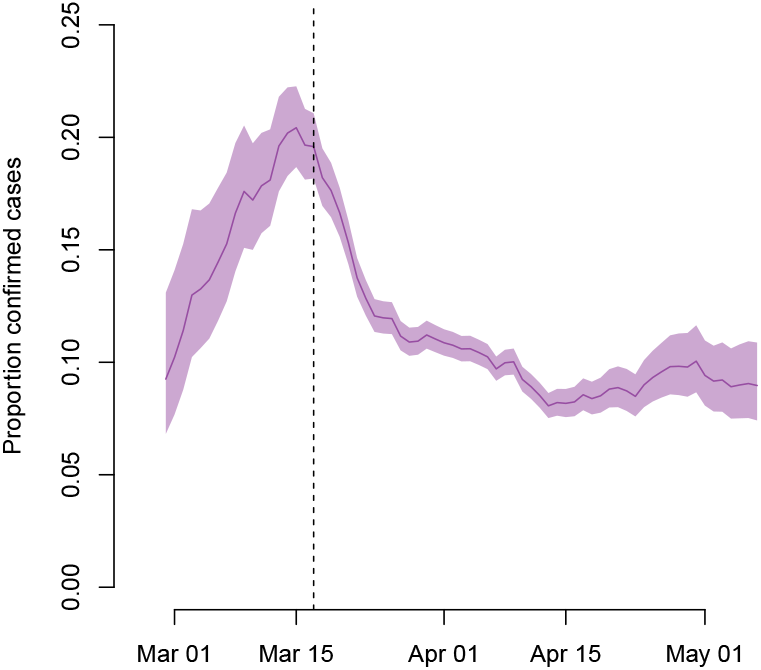
Proportion of new SARS-CoV-2 infections that were diagnosed and reported. The solid line represents the moving average (7 days) of the ratio of confirmed cases over the total number of new infections, and the shaded area corresponds to the 95% prediction interval. The proportion of reported infections increased during the early phase of the epidemic and decreased again during the ‘lockdown’. We assumed that confirmed infections get reported 6 days after onset of symptoms. The vertical dashed line indicates the strengthening of social distancing on 17 March 2020.

We simulated counterfactual scenarios of the SARS-CoV-2 epidemic in Switzerland with an earlier and later initiation of the consecutive introduction of NPIs (table 2). In the baseline scenario (0 days), the model estimated that 2,003 (95% PI: 1,875–2,125) deaths would have occurred assuming NPIs continued to reduce the number of infections after 10 May 2020. Due to the rapid exponential growth of the epidemic at the end of February and in early March 2020, implementing the same sequence of NPIs one week earlier would have resulted in a considerably lower number of peak hospitalization (439, 95% PI: 367–517), peak ICU occupancy (88, 95% PI: 53–123) and deaths (399, 95% PI: 347–458). In contrast, implementing the NPIs one week later, the numbers of peak hospitalization (10,198, 95% PI: 9,245–11,147), peak ICU occupancy (1,908, 95% PI: 1,685–2,149) and deaths (8,683, 95% PI: 8,038–9,453) would have increased substantially.

**Table 2.** Counterfactual scenarios for the SARS-CoV-2 epidemic in Switzerland from February to May 2020.^a^.

| Implementation of NPIs | Maximal number of daily infections | Total infections<sup>b</sup> | Maximal number of hospitalized patients | Maximal number in ICU | Maximal number of daily deaths | Total deaths<sup>b</sup> |
| --- | --- | --- | --- | --- | --- | --- |
| -7 days | 2,827 (2,610–3,061) | 53,278 (48,266–58,591) | 439 (367–517) | 88 (53–123) | 16 (13–21) | 399 (347–458) |
| -6 days | 3,566 (3,331–3,844) | 67,220 (61,594–73,250) | 553 (473–641) | 109 (70–152) | 20 (16–25) | 505 (446–574) |
| -5 days | 4,508 (4,223–4,816) | 84,908 (78,477–91,550) | 701 (611–793) | 137 (94–182) | 24 (20–30) | 637 (570–705) |
| -4 days | 5,696 (5,361–6,034) | 106,982 (99,861–114,421) | 882 (777–993) | 170 (118–221) | 30 (25–36) | 803 (726–879) |
| -3 days | 7,195 (6,761–7,613) | 134,775 (126,931–143,208) | 1,112 (998–1,242) | 216 (161–273) | 36 (31–43) | 1,012 (924–1,098) |
| -2 days | 9,072 (8,538–9,627) | 169,470 (160,894–178,904) | 1,406 (1,276–1,554) | 268 (207–335) | 44 (38–53) | 1,271 (1,181–1,364) |
| -1 days | 11,435 (10,757–12,180) | 212,820 (202,619–223,389) | 1,763 (1,612–1,927) | 337 (264–413) | 54 (47–64) | 1,597 (1,484–1,708) |
| 0 days | 14,404 (13,486–15,383) | 266,417 (254,533–279,072) | 2,213 (2,044–2,388) | 423 (341–501) | 67 (59–77) | 2,003 (1,875–2,125) |
| 1 days | 18,135 (16,924–19,496) | 332,682 (319,241–348,743) | 2,776 (2,586–2,983) | 527 (440–615) | 82 (74–96) | 2,499 (2,358–2,647) |
| 2 days | 22,754 (21,167–24,726) | 414,519 (396,324–436,283) | 3,484 (3,223–3,733) | 657 (556–763) | 101 (90–116) | 3,106 (2,940–3,293) |
| 3 days | 28,512 (26,325–31,244) | 514,168 (490,070–545,328) | 4,345 (4,027–4,673) | 825 (704–947) | 125 (111–141) | 3,861 (3,636–4,114) |
| 4 days | 35,588 (32,692–39,483) | 634,745 (601,950–676,270) | 5,414 (5,025–5,860) | 1,020 (881–1,156) | 153 (138–172) | 4,765 (4,479–5,110) |
| 5 days | 44,428 (40,382–49,535) | 780,140 (734,854–840,806) | 6,720 (6,193–7,266) | 1,254 (1,102–1,424) | 188 (168–210) | 5,852 (5,477–6,323) |
| 6 days | 55,119 (49,904–62,132) | 952,888 (892,386–1,031,927) | 8,288 (7,598–9,055) | 1,556 (1,359–1,771) | 229 (205–257) | 7,150 (6,662–7,772) |
| 7 days | 68,143 (61,154–77,341) | 1,155,026 (1,077,718–1,258,265) | 10,198 (9,245–11,147) | 1,908 (1,685–2,149) | 278 (248–315) | 8,683 (8,038–9,453) |

<sup>a</sup> Numbers represent medians and 95% prediction intervals in brackets<sup>b</sup> Includes infections and deaths assuming NPIs continued after 10 May 2020

## DISCUSSION

Using a dynamic transmission model, we described the successful containment of the SARS-CoV-2 epidemic in Switzerland from February to May 2020. We estimated that the epidemic initially expanded with an *R*_0_ = 2.61 (95% CI: 2.51–2.71) and that the implementation of NPIs reduced transmission to *R*_*e*_ = 0.64 (95% CI: 0.61–0.66). We showed that the proportion of SARS-CoV-2 infections that were diagnosed and reported increased during the early phase of the epidemic and decreased to around 10% after implementation of all NPIs. The counterfactual scenarios illustrate that introducing NPIs one week earlier or later would have resulted in 399 (95% PI: 347–458) and 8,683 (95% PI: 8,038–9,453) deaths, respectively. The urgency of a rapid implementation of NPIs is further underlined by our finding that one week of uncontrolled spread of SARS-CoV-2 in Switzerland required 3.1 weeks (95% CI: 2.8–3.3 weeks) of ‘lockdown’.

A major strength of our study is that our model relies on publicly available data on the number of hospitalized cases, ICU occupancy, and deaths. This allowed us to infer the expected number of new SARS-CoV-2 infections and estimate the level of underreporting of infections during the epidemic in Switzerland. The transmission model is informed using literature values and we used a maximum likelihood framework to estimate critical parameters and their associated uncertainty that describe the epidemic in Switzerland. To our knowledge, our study is the first to explore the potential consequences of counterfactual scenarios on the timing of implementation of NPIs.

Our study comes with a number of limitations. First, we did not have access to line lists of patients and used aggregated data of SARS-CoV-2 cases. Compared to the numbers communicated by the Swiss Federal Office of Public Health (FOPH), cantons typically report somewhat higher numbers of hospitalized and deceased patients. For example, FOPH communicated only 1,640 deaths [1], while the cantonal data reported 1,860 deaths by 10 May 2020. The discrepancies may result from missing or delayed patient records that cannot be linked in the line lists at FOPH. Second, we opted for a maximum likelihood framework and fixed a number of model parameters, such as hospitalization periods, to values that were informed by the literature. While our model can accurately describe the changes in hospitalized patients and ICU occupancy, the calculated CIs can be overly narrow for some parameters. Third, we described the SARS-CoV-2 epidemic in Switzerland overall and did not consider cantonal differences in the transmission dynamics as in Lemaitre et al. [11]. We think that this simplification is justified for the purpose of our study but acknowledge that cantonal differences in the epidemic trajectories were arguably important for the timing of the implementation of NPIs at the federal level. Fourth, we did not stratify the population by sex and age and cannot provide age-specific infection attack rates [24]. Fifth, we assumed a fixed IFR of 0.75% [11], which is in the range of estimates for Switzerland and similarly affected European areas [9, 10]. Higher or lower IFRs would result in lower and higher model estimates of the infection attack rate, and higher and lower estimates of the proportions of SARS-CoV-2 infections that were diagnosed and reported. Sixth, the model parameters on the probability of death in hospital bed (*ε*_3_) and ICU (*ε*_4_) need to be interpreted with caution, as they contribute to a phenomenological description of the hospitalization pathway and implicitly account for deaths outside of hospitals as well. Hence, it is worth noting that the probability of death for ICU patients in Switzerland was reported at 23% [25], which is lower than the 50% used in our model. Finally, our modeling framework was not adapted to investigate the individual effects of single NPIs between 28 February and 20 March 2020. Instead, we assumed that NPIs in Switzerland primarily reduced transmission during a period of two weeks around 17 March 2020, which is in agreement with the study by Lemaitre et al. [11]. Similarly, Flaxman et al. [12] estimated that the majority of reduction in transmission in Switzerland derived from the banning of gatherings of more than five people on 20 March 2020, and Scire et al. [26] showed that the reproduction number dropped significantly below the critical threshold of one after that date.

Our estimates of *R*_0_ and *R*_*e*_ are similar to those found in other studies for Switzerland [11, 26]. The estimated infection attack rate for Switzerland overall is also in line with the study by Lemaitre et al. [11], and in between estimates from seroprevalence studies from low and high incidence cantons, such as Zurich with 1.6% [27] and Geneva with 10.8% [24]. While our estimate of *R*_0_ for Switzerland is similar to early estimates for the epidemic in Wuhan, China [3, 4], we found a faster epidemic spread with a lower doubling time. The estimated doubling time for Switzerland is consistent with the epidemic expansion in other regions of Europe during late February and early March. However, it could also be biased due to underreporting of hospitalized patients and SARS-CoV-2-related deaths during the early phase of the epidemic. A lower epidemic doubling time before the implementation of NPIs in Switzerland would result in smaller differences in peak hospitalization and ICU occupancy and total numbers of deaths between the counterfactual scenarios that are presented.

The results from our study support the notion that an earlier implementation of NPIs in Switzerland would have reduced the number of hospitalizations and deaths due to SARS-CoV-2 considerably. This finding may partially explain the lower numbers of SARS-CoV-2-related deaths in Austria, which was similarly affected by the proximity to the early outbreak in Italy but implemented strict control interventions at an earlier time point compared to Switzerland [12, 28]. Implementing the sequence of NPIs in Switzerland only one week later would likely have led to an overburdening of the healthcare system with a peak demand for ICUs of 1,908 (95% PI: 1,685–2,149), which is higher than the effective *ad-hoc* ICU bed capacity of 1,275 in April 2020 [25].

Our study illustrates that time is of the essence when it comes to outbreak response. Each week of uncontrolled spread of SARS-CoV-2 in Switzerland required roughly three weeks of ‘lockdown’ to reduce the number of new infections to the same level again. There are several lessons to be learnt for the further management of the SARS-CoV-2 epidemic in Switzerland. First, a rapid implementation of NPIs in the case of a resurgence of the epidemic can limit the necessary duration of strict control measures possibly resulting in lower economic and social costs. Second, new epidemic flare-ups are likely to occur in regionally confined areas given the cantonal differences in incidence. A rapid outbreak response at the local level would likely be most effective in such situations. In summary, we showed that while an earlier implementation of NPIs in Switzerland would have resulted in considerably lower numbers of hospitalized cases and deaths, the measures that were taken successfully prevented a much higher burden of SARS-CoV-2-related morbidity and mortality.

## Data Availability

All data and R code files are available on GitHub: https://github.com/calthaus/swiss-covid-epidemic.

